# Comparative analysis of loop-mediated isothermal amplification (LAMP)-based assays for rapid detection of SARS-CoV-2 genes

**DOI:** 10.1101/2020.12.21.20248288

**Authors:** Daniel Urrutia-Cabrera, Roxanne Hsiang-Chi Liou, Jianxiong Chan, Sandy Shen-Chi Hung, Alex W Hewitt, Keith R Martin, Patrick Kwan, Raymond Ching-Bong Wong

## Abstract

The COVID-19 pandemic caused by SARS-CoV-2 has infected millions worldwide and there is an urgent need to increase our diagnostic capacity to identify infected cases. Although RT-qPCR remains the gold standard for SARS-CoV-2 detection, this method requires specialised equipment in a diagnostic laboratory and has a long turn-around time to process the samples. To address this, several groups have recently reported development of loop-mediated isothermal amplification (LAMP) as a simple, low cost and rapid method for SARS-CoV-2 detection. Herein we present a comparative analysis of three LAMP-based assays that target different regions of the SARS-CoV-2: ORF1ab RdRP, ORF1ab nsp3 and Gene N. We perform a detailed assessment of their sensitivity, kinetics and false positive rates for SARS-CoV-2 diagnostics in LAMP or RT-LAMP reactions, using colorimetric or fluorescent detection. Our results independently validate that all three assays can detect SARS-CoV-2 in 30 minutes, with robust accuracy at detecting as little as 1000 RNA copies and the results can be visualised simply by color changes. We also note the shortcomings of these LAMP-based assays, including variable results with shorter reaction time or lower load of SARS-CoV-2, and false positive results in some experimental conditions. Overall for RT-LAMP detection, the ORF1ab RdRP and ORF1ab nsp3 assays have higher sensitivity and faster kinetics for detection, whereas the Gene N assay exhibits no false positives in 30 minutes reaction time. This study provides validation of the performance of LAMP-based assays for SARS-CoV-2 detection, which have important implications in development of point-of-care diagnostic for SARS-CoV-2.

## Introduction

The outbreak of Severe Acute Respiratory Syndrome Coronavirus 2 (SARS-CoV-2) has caused the catastrophic COVID-19 pandemic, which has infected >63 million and caused >1.4 million deaths worldwide to date (Johns Hopkins Coronavirus Resource Centre). Reverse transcription quantitative PCR (RT-qPCR) remained the gold standard diagnostic test for SARS-CoV-2 in many countries, with standard diagnostic protocol from the World Health Organisation. However, this procedure requires processing at a clinical diagnostic laboratory which would take hours or days, depending on the workload at the test site. Given the high infectious rate of SARS-CoV-2 with >500,000 daily new cases globally, there is an urgent need to develop scalable, low cost detection methods for SARS-CoV-2 to increase our capacity to perform daily testing.

Loop-mediated isothermal amplification (LAMP) is a one-step nucleic acid amplification method that has been applied for diagnostic of infectious diseases [1]. The LAMP reaction involves auto-cycling strand displacement DNA synthesis, using hairpin-forming LAMP primers which anneal to the target DNA template and a DNA polymerase with strand displacement activity. These annealed LAMP primers are in turn displaced by displacement primers for subsequent amplification and elongation [2]. The LAMP reaction is highly sensitive, specific and only requires one set temperature for nucleic acid amplification, providing an attractive technology for the development of a low cost, point-of-care diagnostics for SARS-CoV-2. Recently, there has been significant development on the use of LAMP for detection of SARS-CoV-2. Several LAMP assays have been developed that target different regions of SARS-CoV-2, including RNA-dependent RNA polymerase (RdRP) [3], non-structural protein 3 (nsp3) [4] and the nucleocapsid (N) gene [5]. However, how these LAMP assays perform relative to each other remain unclear and understanding this would facilitate development of a robust LAMP assay for SARS-CoV-2 detection.

In this study, we perform a comparative analysis of three of the earliest LAMP assays developed for SARS-CoV-2 detection [3–5]. We perform a head-to-head comparison of the sensitivity and kinetics to detect the RNA or cDNA of SARS-CoV-2 genes, as well as their capability to be used as colorimetric or fluorescent detection.

## Materials and Methods

### Molecular reagents

For positive DNA control, two regions in SARS-CoV-2 ORF1, nt.2945-3370 and nt.14971-15970, were synthesized as gBLOCK double stranded DNA fragments (Integrated DNA Technologies), as well as the 2019-nCoV_N positive control plasmid carrying the SARS-CoV-2 Gene N (GenBank: NC_045512.2, Integrated DNA Technologies).

For positive RNA control, we used the Twist Synthetic SARS-CoV-2 RNA control which provides coverage of >99.9% of the viral genome in six non-overlapping 5kb single stranded RNA fragments (GeneBank: MT007544.1, Twist Bioscience).

### RT-LAMP/LAMP colorimetric assay

Positive DNA or RNA controls were diluted to various concentrations and used in LAMP and RT-LAMP reactions respectively. The sequences of primers and controls used are listed in Supplementary table 1. The six primers (F3, B3, FIP, BIP, Loop F and Loop B) were premixed as a 10X working stock containing 2 µM of each outer primer (F3 & B3), 16 µM of each inner primer (FIP & BIP), and 4 µM of each loop primer (Loop F & Loop B).

For colorimetric detection, a 20 μl reaction was set up containing 10 μl of WarmStart Colorimetric LAMP 2X Master Mix (New England Biolabs), 2 μl primer mix, 1μl RNA/DNA control and 7 μl nuclease-free water. The mixed reactions were incubated at 65°C using a heat block and pictures for colorimetric changes were taken from 10-45 minutes. Subsequently, the amplified samples were run on an 1% agarose gel to determine amplicon specificity.

### RT-LAMP fluorescent assay

Same sets primer and positive RNA controls were used for the RT-LAMP colorimetric assay. For fluorescent detection of RT-LAMP, a 20 μl reaction was set up containing 10μl of WarmStart Colorimetric LAMP 2X Master Mix (New England Biolabs), 2 μl primer mix, 1 μl RNA control, 1 μl of 10X SYBR green (Thermo Fisher) and 7 μl nuclease-free water. The mixed reactions were incubated at 65°C using a StepOne Real Time PCR machine (Thermo Fisher) for 60 minutes and fluorescence measurements were taken every 2.5 minutes.

To quantify the kinetics of the RT-LAMP assay, the ΔRn values are extracted and plotted against the 2.5 timepoints. A threshold corresponding to half the maximum ΔRn value is set to determine the time required to reach half the maximum fluorescence intensity, termed ‘½ ΔRn_max_’. If there is no detected signal after 60 minutes reaction time, the ½ ΔRn_max_ is listed as 60 minutes.

## Results

### Comparative study of LAMP reactions using colorimetric detection

For this study, we used the WarmStart Colorimetric RT-LAMP 2X Master Mix (New England Biolabs) containing a warm-start reverse transcriptase RTx and an isothermal DNA polymerase Bst 2.0, which is capable of both LAMP and RT-LAMP. The reaction mix also contains a pH indicator, which allows visualisation of amplification as a result of protons produced by the LAMP reaction and lead to a red-to-yellow colour change. We selected primer sets reported by three previous studies that targeted different regions of the SARS-CoV-2 genome (Supplementary table 1). The ‘ORF1ab RdRP’ set [3] and the ‘ORF1ab nsp3’ set [4] targeting RdRP and nsp3 sequences encoded by ORF1ab respectively, as well as the ‘Gene N’ set which targets the nucleocapsid gene [5].

We first assessed the capability of these three primer sets in colorimetric LAMP reactions, using synthesized DNA of SARS-CoV-2 genes as control. Our results showed that the three primer sets are all capable of LAMP detection of the SARS-CoV-2 genes, albeit with some differences in performances (Figure 1). In terms of kinetics for the colorimetric LAMP reaction, visible colour changes can be seen as early as 20 minutes for ORF1ab RdRP and Gene N, compared to 25 minutes for ORF1ab nsp3. In order to determine the sensitivity of the LAMP assays, we performed LAMP reactions with a dilution series ranging from 100 to 10000 copies of DNA molecules. Overall the lowest detection limit is 100 copies of DNA molecules for both ORF1ab nsp3 and Gene N, compared to 500 copies of DNA molecules for ORF1ab RdRP (Figure 1). In addition, gel electrophoresis showed a discrete band representing the LAMP amplicons (Figure 2A-C), supporting the specificity of the LAMP amplification for all three primer sets. Collectively, all three primer sets are capable of colorimetric LAMP detection of SARS-CoV-2 within 30 minutes, with slightly faster kinetics for the ORF1ab RdRP/Gene N and slightly higher sensitivity for ORF1ab nsp3.

**Figure 1:**
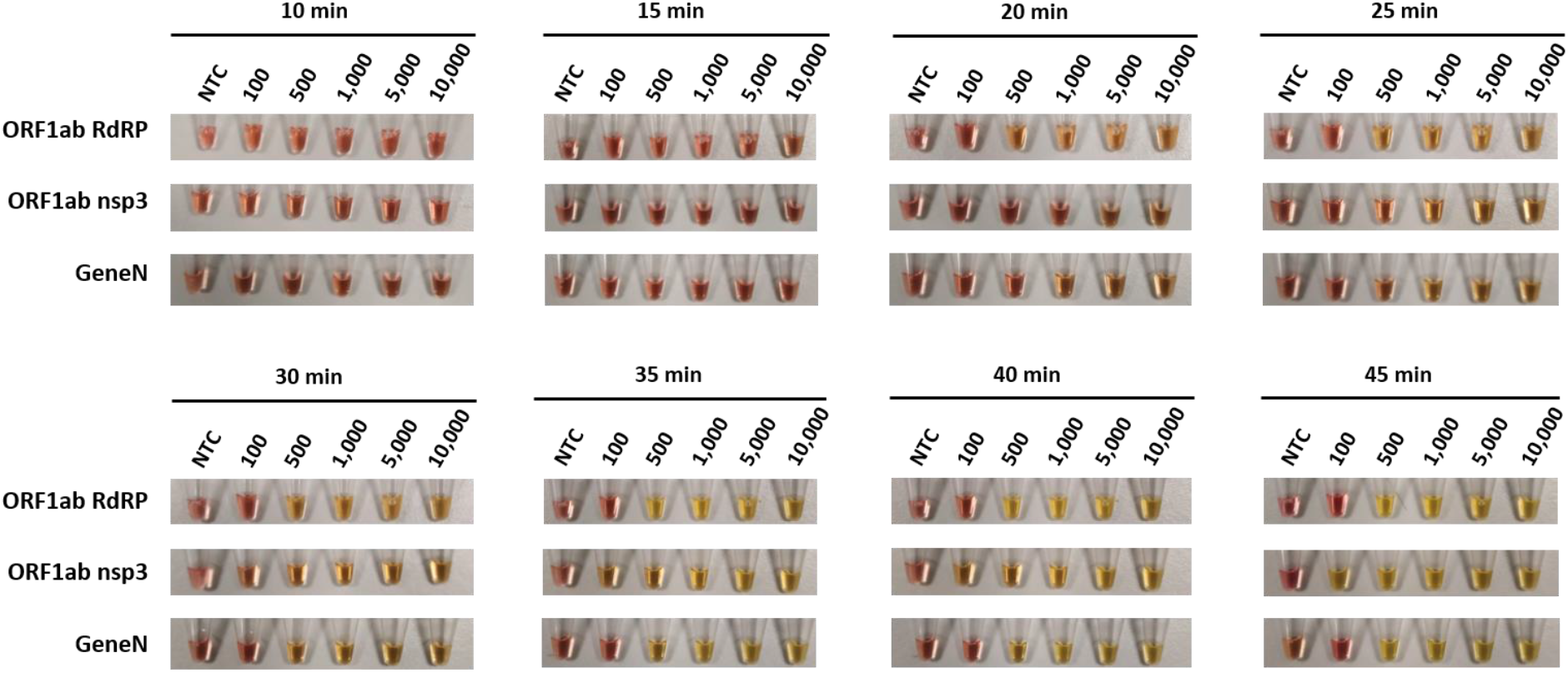
Colorimetric detection of SARS-CoV-2 DNA using LAMP. LAMP reactions are assessed at different time points using a dilution series of positive DNA control ranging from 100 copies to 10000 copies. NTC (no template control) is used as a negative control. n=3 biological repeats.

**Figure 2:**
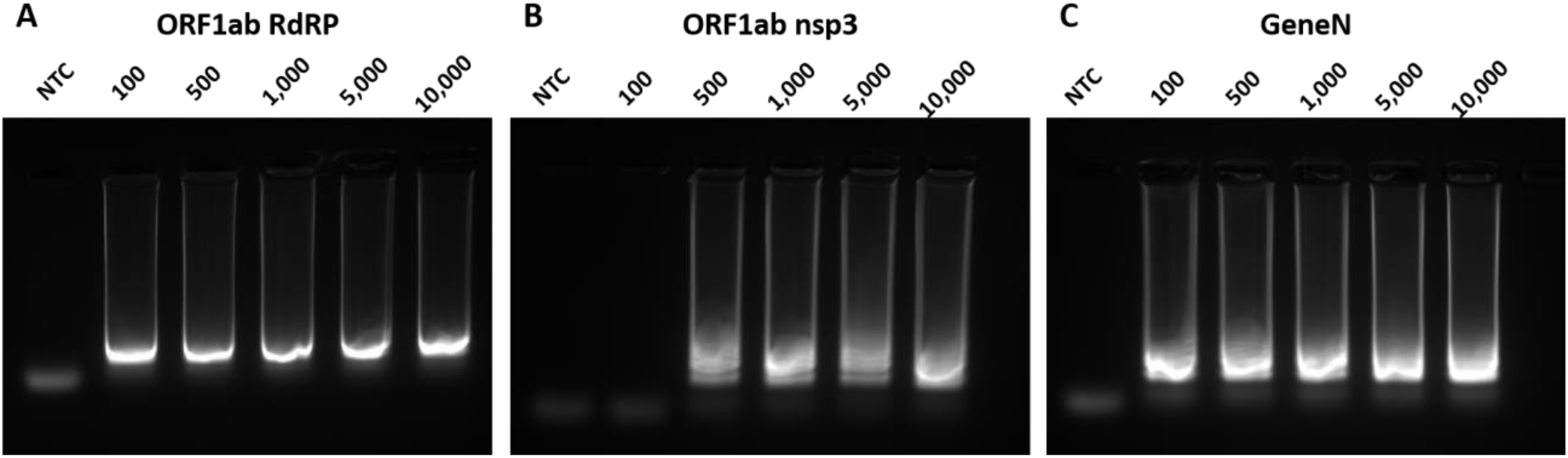
Gel electrophoresis analysis of LAMP reaction. Representative gel picture is shown for A) ORF1ab RdRP, B) ORF1ab nsp3 and C) Gene N in LAMP assays, using a dilution series of positive DNA control ranging from 100 copies to 10000 copies. NTC (no template control) is used as a negative control. n=3 biological repeats.

### Comparative study of RT-LAMP reactions using colorimetric detection

Next, we compared the three primer sets in colorimetric RT-LAMP reactions, using in vitro transcribed RNA of SARS-CoV-2 genes as control. Similar to our LAMP results, the ORF1ab RdRP is the fastest out of the three with visible colour change after 20 minutes of reaction, whereas ORF1ab nsp3 and Gene N take 25 minutes and 30 minutes respectively (Figure 3). In terms of sensitivity, the lowest detection limit is observed in ORF1ab RdRP (100 copies of RNA molecules), followed by ORF1ab nsp3 and Gene N (both 500 copies of RNA molecules, Figure 3). However for the ORF1ab RdRP and nsp3 primer sets, we noticed that the no template control sometimes exhibits colour change indicative of a false-positive, while this was not observed for the Gene N set (Table 1).

**Table 1:** Summary of false positives detected in no template control in individual biological repeats.

| Primer sets | Assay | Total false positive | False positive after 15 min | False positive after 30 min | False positive after 45 min |
| --- | --- | --- | --- | --- | --- |
| ORF1ab RdRP | LAMP | 4/6 (66%) | 0/6 (0%) | 3/6 (50%) | 4/6 (66%) |
|  | RT-LAMP | 4/5 (80%) | 1/5 (20%) | 4/5 (80%) | 4/5 (80%) |
| ORF1ab nsp3 | LAMP | 2/6 (33%) | 0/6 (0%) | 1/6 (16%) | 2/6 (33%) |
|  | RT-LAMP | 3/5 (60%) | 1/5 (20%) | 2/5 (40%) | 3/5 (60%) |
| Gene N | LAMP | 3/6 (50%) | 0/6 (0%) | 0/6 (0%) | 3/6 (50%) |
|  | RT-LAMP | 0/5 (0%) | 0/5 (0%) | 0/5 (0%) | 0/5 (0%) |

**Figure 3:**
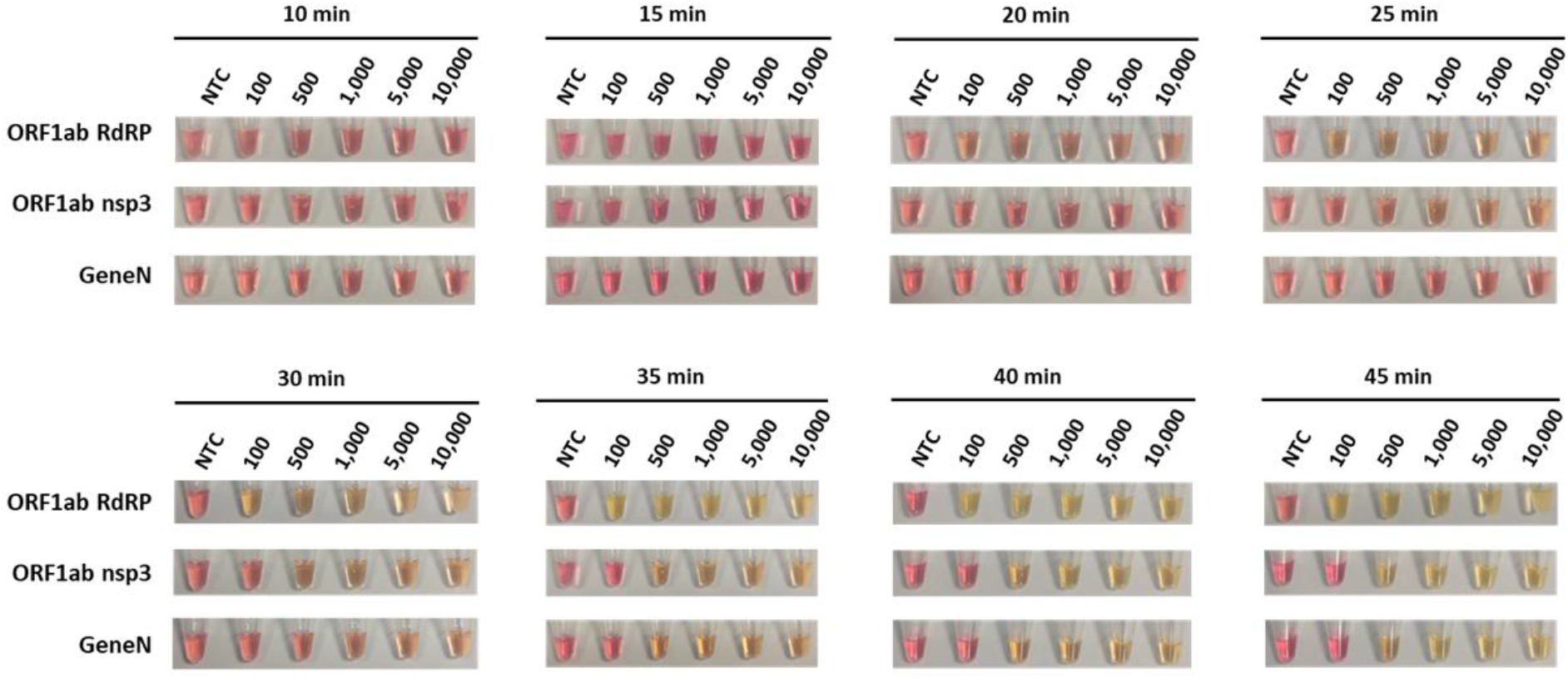
Colorimetric detection of SARS-CoV-2 RNA using RT-LAMP. RT-LAMP reactions are assessed at different time points using a dilution series of positive RNA control ranging from 100 copies to 10000 copies. NTC (no template control) is used as a negative control. n = 3 biological repeats.

Correspondingly, analysis by gel electrophoresis revealed a clear band of the correct RT-LAMP reaction product in lanes with ≥100 copies RNA input for ORF1ab RdRP, and ≥500 copies RNA input for both ORF1ab nsp3 and Gene N (Figure 4A-C). These results support the specificity of the RT-LAMP amplification using the three primer sets. Overall, the three primer sets perform similarly in LAMP or RT-LAMP for colorimetric detection, with only subtle differences in sensitivity and kinetics.

**Figure 4:**
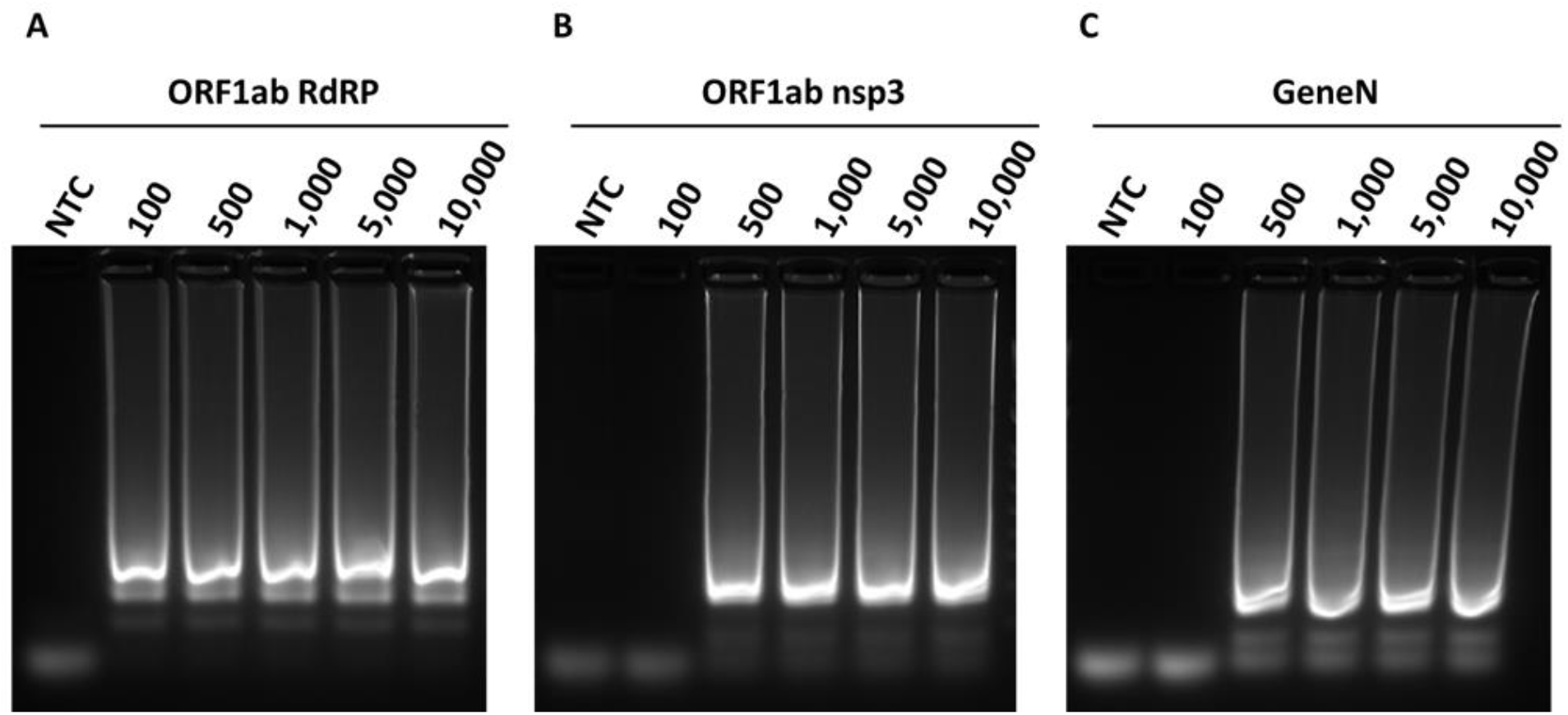
Gel electrophoresis analysis of RT-LAMP reaction. Representative gel picture is shown for A) ORF1ab RdRP, B) ORF1ab nsp3 and C) Gene N in RT-LAMP assays, using a dilution series of positive RNA control ranging from 100 copies to 10000 copies. NTC (no template control) is used as a negative control. n = 3 biological repeats.

### Assessment of the kinetics and sensitivity of RT-LAMP reactions using fluorescent detection

Although colorimetric detection represents an easy detection method for point-of-care diagnostic development, subtle colour changes may not be obvious with naked eye. To more accurately assess the sensitivity and kinetics of the three primer sets, we added SYBR Green I into the RT-LAMP reactions which allows fluorescent quantification using a qPCR machine. For all three primer sets, increase in the fluorescent signal starts around 12-15 minutes. The fluorescent signal reached a plateau around 30-40 minutes for ORF1ab RdRP and ORF1ab nsp3 (Figure 5A, C), indicating that 40 minutes of reaction is sufficient for these two primer sets. However, Gene N exhibits different amplification curves depending on the copy number of the RNA molecules: ∼30 minutes should be sufficient for detection of ≥500 copies, whereas >45 minutes is needed for detection of 100 copies of RNA molecules (Figure 5E).

**Figure 5:**
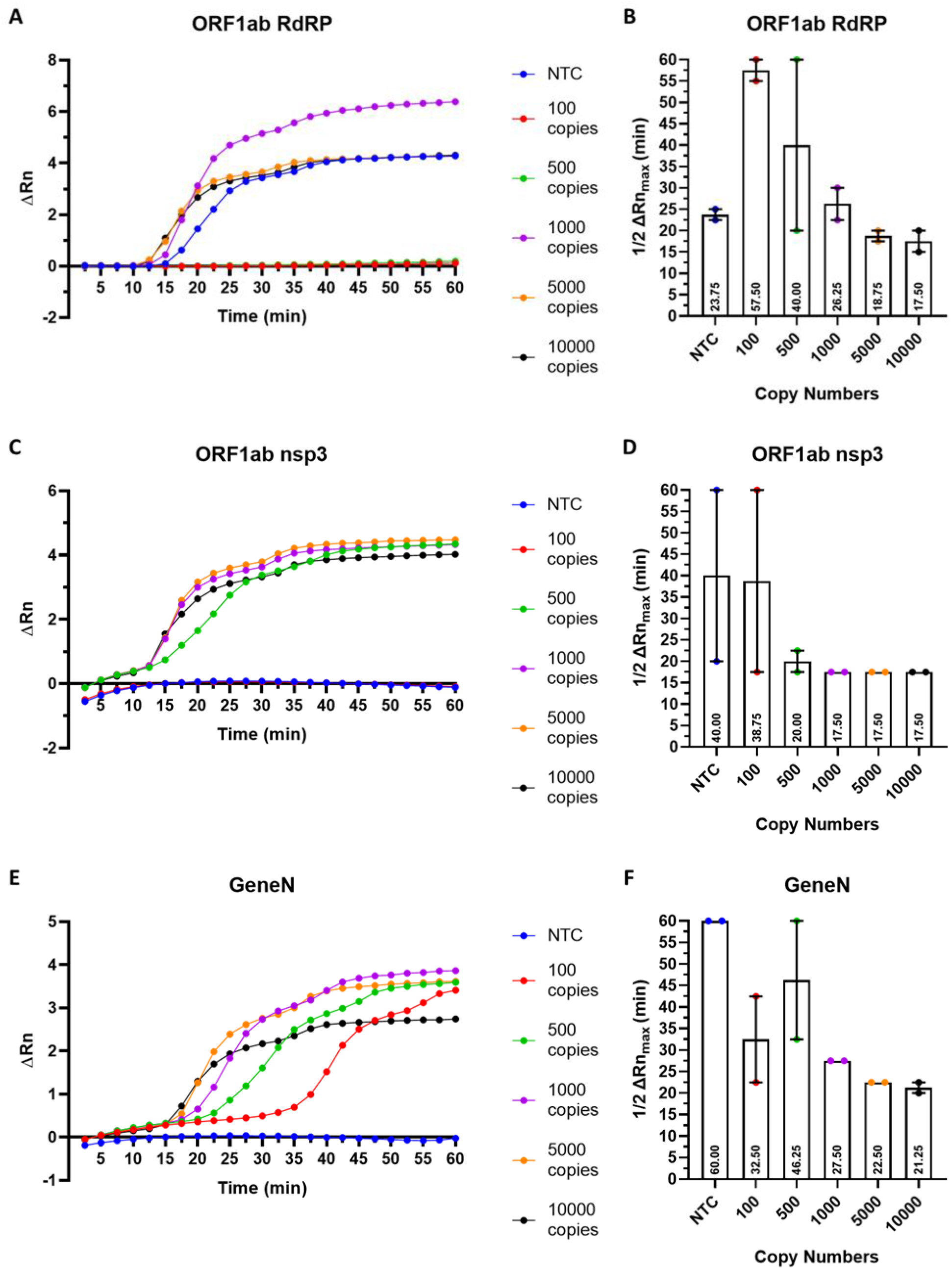
Fluorescent detection of SARS-CoV-2 RNA using RT-LAMP. Real-time measurement of SARS-CoV-2 RNA using RT-LAMP for A) ORF1ab RdRP, C) ORF1ab nsp3 and E) Gene N, showing the normalised fluorescent signal (ΔRn) against the reaction time. Quantification of the time to reach half the maximum fluorescent signal (ΔRn_max_) for B) ORF1ab RdRP, D) ORF1ab nsp3 and F) Gene N. Results are displayed as the mean of 2 biological repeats, error bars represented SEM.

To compare the kinetics of RT-LAMP across the three primer sets, we measure the time required to reach half the maximum fluorescence signal (½ ΔRn_max_). We noted that ½ ΔRn_max_ is generally consistent for detection of ≥1000 copies of RNA molecules, whereas detection of 100 or 500 copies of RNA molecules yielded variable ½ ΔRn_max_values in biological repeats. This suggested the three primer sets perform more robustly for detection of higher copies of SARS-CoV-2 RNA. For detection of 1000 copies of RNA molecules, ORF1ab nsp3 exhibit the fastest kinetics (½ ΔRn_max_ = 17.5 min), followed by ORF1ab RdRP (½ ΔRn_max_ = 26.25 min) and Gene N (½ ΔRn_max_ = 27.5 min). However, for detection of 10000 copies of RNA molecules, all three primer sets exhibit comparable kinetics (½ ΔRn_max_ = 17.5 - 21.25 min) (Figure 5B, D, F). Collectively, our results suggest that the three RT-LAMP assays have comparable kinetics for detection of high copies of SARS-CoV-2, but variable performances for detection of low copies of SARS-CoV-2.

### False positive rates for RT-LAMP/LAMP reactions

In our testing, occasionally we detected positive signals in the no template controls which are indicative of false positives. Table 1 lists the false positive rate for the three primer sets in RT-LAMP and LAMP reactions. In general, we observed lower false positive rates in shorter reaction time (15 minutes) and higher false positive rates in longer reaction time (45 minutes), thus we recommend using a reaction time of 30 minutes. Notably in a 30 minutes RT-LAMP reaction, the lowest false positive rate is observed in Gene N (0/5, 0%), followed by ORF1ab nsp3 (2/5, 40%) and ORF1ab RdRP (4/5, 80%). These results suggest that the Gene N primer set has the best accuracy for detection among the three tested assays.

## Discussion

Active cases of SARS-CoV-2 can be diagnosed by detection of the viral RNA or antigens in patient samples. While antigen-based detection methods are cheap and rapid as a point-of-care diagnostics, they are inherently less sensitive compared to viral RNA detection methods given there is no amplification of protein signals. As such, the sensitivity of many antigen-based detection methods ranges from 50-90% [6]. In contrast, viral RNA detection methods, such as RT-qPCR remain the gold standard for SARS-CoV-2 diagnostics with ∼98% sensitivity. However, the requirement of sample processing in a clinical diagnostic laboratory results in long turnaround time for diagnostic results and this remained the key bottleneck to upscale our capability to perform daily SARS-CoV-2 testing.

Recent development of RT-LAMP assays could address this key issue, providing a rapid and low-cost detection method for SARS-CoV-2. In this study, we compared and validated three LAMP-based assays previously developed to detect different parts of SARS-CoV-2: ORF1ab RdRP, ORF1ab nsp3 and Gene N. Our results show that the three RT-LAMP assays allow colorimetric detection of SARS-CoV-2 genes in 30 minutes, with robust accuracy at detecting 1000 RNA copies. Also, the three assays perform similarly for detection of RNA or DNA copies of SARS-CoV-2. However, we observed some variability in these RT-LAMP assays with shorter reaction time and detection of lower load of SARS-CoV-2 RNA. Also, in our testing we observed false positive results in some experimental repeats. Overall for RT-LAMP, our comparative analysis showed that the ORF1ab RdRP and ORF1ab nsp3 sets have higher sensitivity and faster kinetics for detection, whereas the Gene N primer set exhibits no false positives in 30 minutes reaction time. Future optimisation of the primer sets and modification of RT-LAMP reactions would be important to develop an accurate and sensitive detection method for SARS-CoV-2, such as the use of quenched fluorescent primers to reduce the false positive rates of RT-LAMP [7]. There are also ongoing development of new RT-LAMP-based assays to improve performance of SARS-CoV-2 detection, including the use of a two-stage isothermal amplifications (Penn-RAMP)[8], integration of CRISPR with RT-LAMP [9,10], as well as barcoded RT-LAMP reactions to allow high-throughput processing using next-generation sequencing (LAMP-seq) [11]. Future validation of these new RT-LAMP-based assays would be important to translate these technologies into clinical testing in the real world.

Although false positives remain an issue, there are several key advantages of the RT-LAMP detection methods. The major advantage is the rapid reaction time of ∼30 minutes, which is far superior compared to RT-qPCR with a turn-around time of several hours to days. Secondly, the readout of RT-LAMP is simple and can be interpreted easily by naked eye, either as a change in colour (this study) or turbidity [12]. Finally, RT-LAMP only requires one isothermal step which makes it simple to develop low cost, portable devices to process the samples. Recently, several companies have developed portable devices to carry out point-of-care RT-LAMP testing, such as DNAfit LifeSciences (UK) and Lucira Health (USA). Moreover, a recent study also demonstrated that RT-LAMP can be successfully conducted in a kitchen range oven, providing a low-cost solution to carry out RT-LAMP testing at home [13]. Further improvement to RT-LAMP would potentially provide an affordable point-of-care detection method for routine SARS-CoV-2 detection.

## Conclusions

In summary, this study shows a detailed assessment of the performances of LAMP-based assays for detection of SARS-CoV-2, providing important validation data that would facilitate further development of LAMP-based diagnostics for SARS-CoV-2 and translation into real world testing.

## Data Availability

Data can be accessed by contacting the corresponding author.

## Abbreviations

SARS-CoV-2: Severe Acute Respiratory Syndrome Coronavirus 2
LAMP: loop-mediated isothermal amplification
RT-LAMP: Reverse transcription loop-mediated isothermal amplification
RT-qPCR: Reverse transcription quantitative PCR
RdRP: RNA-dependent RNA polymerase
nsp3: non-structural protein 3
Gene N: Nucleocapsid gene.

## Declarations

### Ethics approval and consent to participate

Not applicable

### Consent to publication

Not applicable

### Availability of data and materials

The datasets in the current study are available from the corresponding author on reasonable request.

### Authors’ contributions

RCBW, JC and PK designed the experiments; DUC, RHCL conducted the experiments; DUC, RHCL, RCBW, SH, AWH, KM analysed the data; SH, AH, KM, RCBW provided funding to this work; DUC, RHCL and RCBW wrote the manuscript. All authors approved the manuscript.

## Acknowledgements

Not applicable.

---

**Supplementary table 1:**
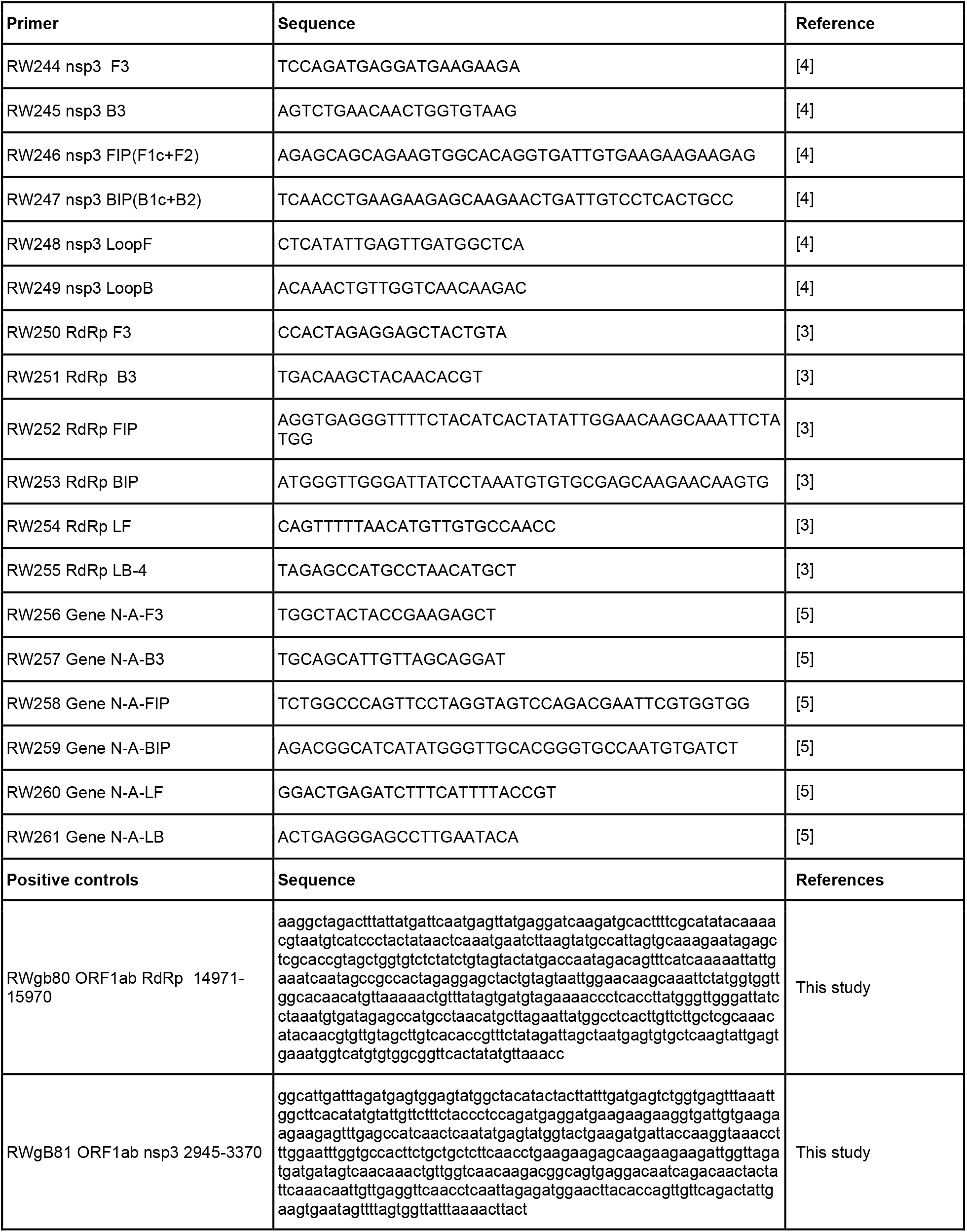
Information of primers and control gBLOCK used in this study.

